# Thermoacoustic ultrasound assessment of liver steatosis - a novel approach for MASLD diagnosis

**DOI:** 10.64898/2025.12.12.25342082

**Authors:** Jang Hwan Cho, Christopher M. Bull, Michael Thornton, Jing Gao, Jonathan M. Rubin, Idan Steinberg

## Abstract

**Background/Objectives:** Metabolic Dysfunction-Associated Steatotic Liver Disease (MASLD) is a global health crisis, but current diagnostics are limited. Liver biopsy is invasive, magnetic resonance imaging-proton density fat fraction (MRI-PDFF) is expensive, and quantitative ultrasound methods are low-accuracy, especially in patients with a high body mass index (BMI). This study introduces a novel thermo-acoustic (TA) method that generates ultrasound signals based on tissue electrical conductivity, where lean tissue (high in water and electrolytes) absorbs more radio-frequency (RF) energy than fatty tissue, providing a direct molecular contrast for fat.

**Methods:** A prospective, cross-sectional feasibility study compared a new thermo-acoustic fat fraction (TAFF) score with the reference standard MRI-PDFF in 40 subjects with suspected fatty liver disease. Bland Altman analysis, Deming regression, and Binary classification performance were tested. To establish system stability, a dedicated Repeatability and Reproducibility (R&R) study (N = 14) evaluated inter-operator and intra-operator consistency using an Intraclass Correlation Coefficient (ICC) derived from a two-way random-effects ANOVA model.

**Results:** TAFF estimates demonstrated a substantial correlation (r =0.89) with MRI-PDFF and an average absolute error of 3.04% fat fraction. Classification performance was high, with an Area Under the Receiver Operating Characteristic Curve (AUROC) of 0.92 at the 12% fat fraction threshold and 0.99 at the 20% fat fraction threshold. The R&R study confirmed robust stability (intraclass correlation = 0.89) and a negligible mean inter-operator difference of 0.36%. Estimation errors showed no statistically significant correlation with BMI or other body habitus measurements.

**Conclusions:** These findings support thermoacoustics’ potential as an accurate, non-invasive, point-of-care solution that can serve as a new imaging biomarker. By providing predictive values closely aligned with MRI-PDFF across the full MASLD spectrum, TAFF can complement currently available ultrasound methods to address the cost and access constraints of MRI for the assessment, diagnosis, and monitoring of MASLD.

## 1. Introduction

### Clinical background

Metabolically dysfunction-associated steatotic liver disease (MASLD), formerly known as nonalcoholic fatty liver disease (NAFLD), is a complex, multi-system metabolic disorder characterized by elevated liver fat fraction (LFF). The steatotic liver is not passive; it acts as an active endocrine organ, secreting hepatokines such as Fetuin-A that drive systemic insulin resistance and chronic inflammation through a vicious cycle of lipotoxicity and oxidative stress [1–11]. This metabolic cascade has profound systemic implications, such as cardiovascular disease (CVD) [12], type 2 diabetes (T2D) [13], hypertension, and neurodegenerative diseases [14]. Recently, MASLD has emerged as a major global health concern, with an estimated prevalence of approximately 30%, establishing MASLD as the most common chronic liver disease in Western nations [15–17].

An estimated 20-30% of individuals with MASLD develop metabolic dysfunction-associated steatohepatitis (MASH), characterized by inflammation and fibrosis that places them at high risk for severe morbidity [18]. This disease progression is clinically significant as MASH is a leading driver for the development of cirrhosis, hepatocellular carcinoma (HCC), liver failure, and ultimately liver transplantation [19,20]. Furthermore, MASLD is strongly associated with a complex of chronic diseases that extend beyond the liver, with CVD representing the primary cause of mortality in the MASLD patient population [12]. However, this risk is dynamic rather than static, as clinical evidence shows that reversing hepatic steatosis successfully ameliorates systemic dysfunction and significantly reduces the risk of associated cardiovascular events [21–25].

Historically, the management of MASH has been predicated upon lifestyle modifications, primarily focused on inducing sustained weight loss [26]. The therapeutic landscape, however, is undergoing a rapid and profound transformation. This evolution was dramatically underscored by the 2024 landmark FDA approval of the first MASH-specific pharmacotherapy, Resmetirom (being marketed as Rezdiffra) [27]. In parallel, robust Phase 3 clinical trial data have demonstrated the compelling efficacy of various incretin therapies, including Glucagon-like Peptide-1 receptor agonists (GLP-1 RAs) such as semaglutide. These agents exhibit high rates of MASH resolution and marked improvements in hepatic fibrosis, firmly establishing them as a cornerstone of future metabolic-based therapeutic strategies [28]. And because MASLD patients die primarily from CVD, GLP-1 RAs offer crucial extrahepatic benefits that target these metabolic complications. Patients with MASLD and coexisting type 2 diabetes, GLP-1 RAs are associated with a significantly lower incidence of major adverse cardiovascular events and all-cause mortality compared to other glucose-lowering drugs [29–31].

Current diagnostic methods for steatotic liver disease exhibit a clear hierarchy based on their accuracy and clinical application. Liver biopsy remains the only definitive method for diagnosing MASH by identifying characteristic inflammation and ballooning; however, its utility for precise, longitudinal quantification is significantly limited by its invasiveness, substantial cost, high risk of sampling error, and inherent inter-observer variability [32,33]. In contrast, magnetic resonance imaging-proton density fat fraction (MRI-PDFF) has emerged as the non-invasive reference standard for quantifying hepatic fat, demonstrating high accuracy and reproducibility, thereby establishing it as the preferred modality for clinical trials [34,35]. Positioning itself as a widely accessible screening method, the first-generation quantitative ultrasound tool, the controlled attenuation parameter (CAP), offers adequate diagnostic accuracy. However, its performance is known to be confounded by factors such as high body mass index (BMI) [36–38]. This emphasis on objective measurement is mandated by the 2025 American Association for the Study of Liver Diseases (AASLD) guidance, which explicitly recommends quantitative methodologies, such as CAP or MRI-PDFF, over standard, subjective gray-scale ultrasound for detecting steatosis [39].

### MASLD diagnosis

The most recent advancements involve state-of-the-art Quantitative Ultrasound (QUS) techniques that leverage advanced analysis of beamformed images or raw radiofrequency ultrasound signals to assess complementary acoustic properties [40–42]. Quantitative ultrasound methods, specifically Attenuation Coefficient (AC) algorithms and/or backscatter signals for liver fat assessment, such as the CAP, ultrasound-derived fat fraction (UDFF), and ultrasound-guided attenuation parameter (UGAP), offer non-invasive approaches for assessing hepatic steatosis.

While the prevalence and related morbidity of MASLD, in combination with the introduction of new treatment options such as GLP-1 drugs, necessitate screening and monitoring of a broad population [43], none of the abovementioned methods is suitable for such a task. Invasive biopsies are painful, costly, and introduce a non-negligible risk to patients [32]. Although MRI-PDFF is accurate and non-invasive [34], it is cost-prohibitive and cannot be performed at the bedside. Ultrasound-based methods, albeit low cost and portable, often suffer from low predictive power, especially in patients with high BMI or confounding fibrosis due to reliance on surrogate parameters in place of fatty tissue molecular contrast. Thus, there is an unmet clinical need for a non-invasive, low-cost, rapid, and portable method with high predictive power for screening and monitoring MASLD treatment. Thermoacoustic (TA) imaging is a hybrid electromagnetic (EM) and acoustic imaging modality that leverages differential absorption of time-varying EM radiation to generate pressure waves in tissue [44]. As EM radiation propagates through tissue, its energy is differentially absorbed by regions based on electrical conductivity [45–47]. This selective absorption results in a rapid, localized increase in temperature. Although the temperature rise is minuscule - often on the order of milli-Kelvins - the almost instantaneous, localized change in tissue temperature is sufficient to generate an acoustic pressure wave that is detectable by an ultrasound transducer array. The wave intensity is proportional to the tissue conductivity, thus providing molecular contrast [48]. The detected pressure signals can then be used to reconstruct images or volumes of data related to tissue composition [49].

Currently, most academic research on TA focuses on system engineering [50–52] or pre-clinical imaging, such as functional brain imaging and cerebral hemorrhage detection [53] or joint and bone imaging [54]. Other examples include deep-tissue (>6 cm) kidney imaging in large swine and real-time monitoring of therapeutic interventions, such as microwave ablation (MWA) therapy [55]. However, in recent years, TA imaging has emerged as a clinical tool for diagnosing and monitoring various tissues and disease states. For example, in their seminal paper, Kruger et al. [56] demonstrated clear contrast enhancement in the tumor region for three patients imaged both before and after chemotherapy. Conversely, the two breast cancer patients imaged after completing their chemotherapy showed no contrast enhancement. This finding correlated with subsequent pathologic examinations, which confirmed their disease was in complete remission.

This paper describes a system and the clinical study used to assess the feasibility of a point-of-care, portable thermoacoustic system for measuring liver fat fraction. To that end, TA techniques can estimate liver fat since their contrast mechanism relies on differences in tissue conductivity when subjected to an RF pulse: lean tissues, which have high water and ion content, absorb significantly more energy than fatty tissues, which have low water and ion content. This difference in energy absorption, particularly at the boundary between muscle and liver, generates a stronger thermoacoustic signal as liver fat content increases, thereby enabling quantitative measurement of steatosis. Moreover, the layered structure of the tissue (skin, subcutaneous fat, muscle, and then liver) provides an inherent calibration method for the measured signal, as subcutaneous fatty tissue is predominantly fat (>90%) and muscle is predominantly lean tissue (>95%). Thus, for each measurement, signals from purely fatty and purely lean tissues are acquired along with those from the unknown liver tissue, enabling their use for calibration. The aim of this paper is to estimate the feasibility, accuracy compared to the reference standard MRI-PDFF, and reproducibility of using a handheld TA device for liver fat estimation.

## 2. Materials and Methods

### TAEUS^®^ Liver system

The TAEUS^®^ Liver System (Endra Life Sciences, Inc., Ann Arbor, Michigan, USA) is a point-of-care thermoacoustic system for measuring LFF. It includes four main subsystems, as illustrated in Figure 1A: (1) Console: A cart-mounted enclosure that contains an RF Source, electronics, and software. (2) Probe: A handheld component that includes an RF applicator and a receive-only ultrasound transducer. (3) Display assembly: A touchscreen tablet that is the graphical user interface for the ultrasound probe, a touchscreen monitor (TA display) that is the graphical user interface and input device. (4) Handheld ultrasound probe: a wireless U/S probe. Figure 1B shows a photo of the system.

**Figure 1.**
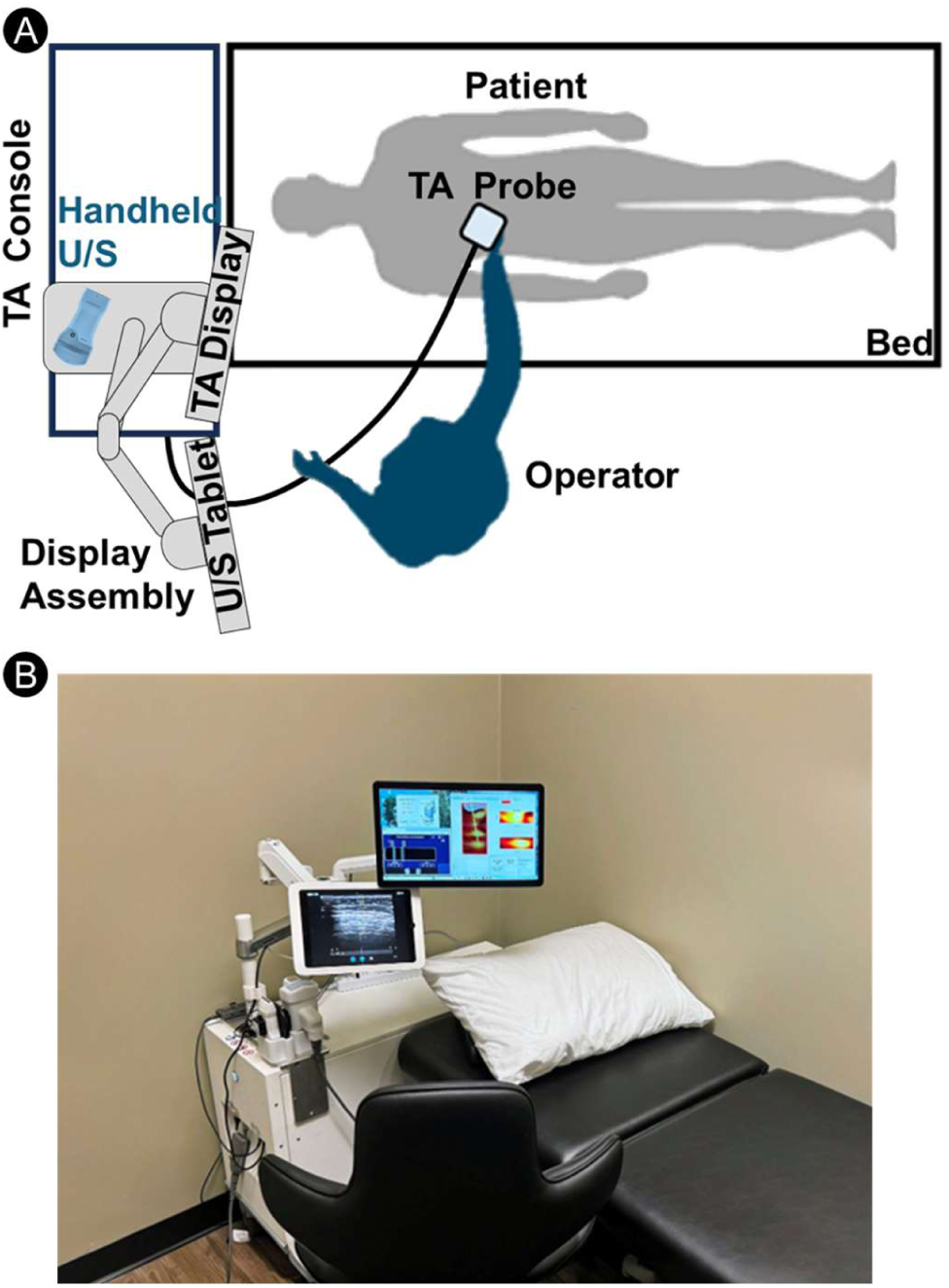
The TAEUS Liver system (A) Illustration of the main subsystems: (A.1) TA console, (A.2) TA probe, (A.3) Display assembly, (A.4) Wireless ultrasound probe. (B) a photo of the scanning room with the TAEUS Liver system

During a scan, the TA console generates high-power RF pulses (7 kW peak power). It delivers these pulses to the RF applicator within the TA probe via a coaxial cable. The RF applicator is an open-ended waveguide antenna designed to efficiently couple RF energy into human tissue. Using an RF tap, a small fraction (<0.01%) of the power is redirected to a power monitor inside the console, ensuring every pulse is monitored for safety. The system operates below the specific absorption rate (SAR) limits for standard mode MRI operation [57] and below the maximal permissible exposure (MPE) for the electric field. [58].

The transducer within the probe receives the ultrasound signal generated by the TA conversion of the RF pulse energy into acoustic energy. The ultrasound signal is then amplified, digitized, and sent to the console for processing and subsequent display on the graphical user interface. A TA exam includes a few preview scans (i.e., 125 pulses each) to ensure that the probe is adequately positioned and can pick a clear liver signal, followed by a longer one or more measurement scans (250 pulses each). Both the preview and measure scans are processed in the same way. Acquired scan data are averaged and digitally filtered to reduce noise and artifacts. A Hilbert transform is applied to the filtered data, followed by universal back-projection [59] and envelope detection to generate the beam-formed image. Since the probe is handheld, its positioning and placement affect the quality of the acquired scan. To identify valid scans with features critical to the accurate thermo-acoustic fat fraction (TAFF) estimation, a scan-quality metric was designed and calculated for each scan. The scan quality metric comprises various geometric and image-based features to ensure that signals are free of interference and their locations are consistent with ultrasound imaging. Measurements with low scan quality were automatically removed from the study. On average, each patient had 5 valid measurements, with a range of 1 to 16. If multiple TAFF scores were obtained per patient, the mean of all TAFF scores was used in the analysis.

For each scan, the TAFF value is estimated from the corresponding beamformed image. Since the TA probe was positioned to include the skin, subcutaneous fat, muscle, and liver in the field of view, the beamformed image from that location is expected to show three signals: the skin-to-subcutaneous fat boundary, the fat-to-muscle boundary, and the muscle-to-liver boundary. Based on these expected signals at the tissue boundaries, the TAFF estimation is formulated as an optimization problem that seeks the liver fat content by minimizing the difference between measured and theoretically expected signal strengths. The cost function relies on a thermoacoustic signal-generation model that incorporates various tissue properties, including the Grüneisen parameter, tissue permittivity, tissue electrical conductivity, and subject-specific electric-field distributions generated by an electric-field model, as described in Appendix A. The electric field model generated the necessary electric field distributions for subjects with various body habitus and liver fat content, and was estimated using Ansys HFSS (Ansys, Canonsburg, PA, USA) simulation software to account for these variations.

### Study description

To assess the ability of TA to accurately estimate LFF across various disease states and body habitus, a prospective, HIPAA-compliant, cross-sectional study was conducted as a proof-of-concept and preliminary evaluation of the method. A single-site (Ann Arbor, MI, USA) single-operator study was approved by the Advarra Institutional Review Board (IRB approval number: Pro00089043). First, prospective participants recruited from the local population were screened via questionnaire for clinically proven or suspected elevated liver steatosis (based on known comorbidities, lifestyle, and body habitus). If liver steatosis was confirmed or suspected, participants were invited to join the study, and written informed consent was obtained. This was done primarily to increase the number of subjects with liver steatosis in the study and to enable testing of the system across all disease grades. Inclusion and exclusion criteria are described in Table 1 below:

**Table 1.**
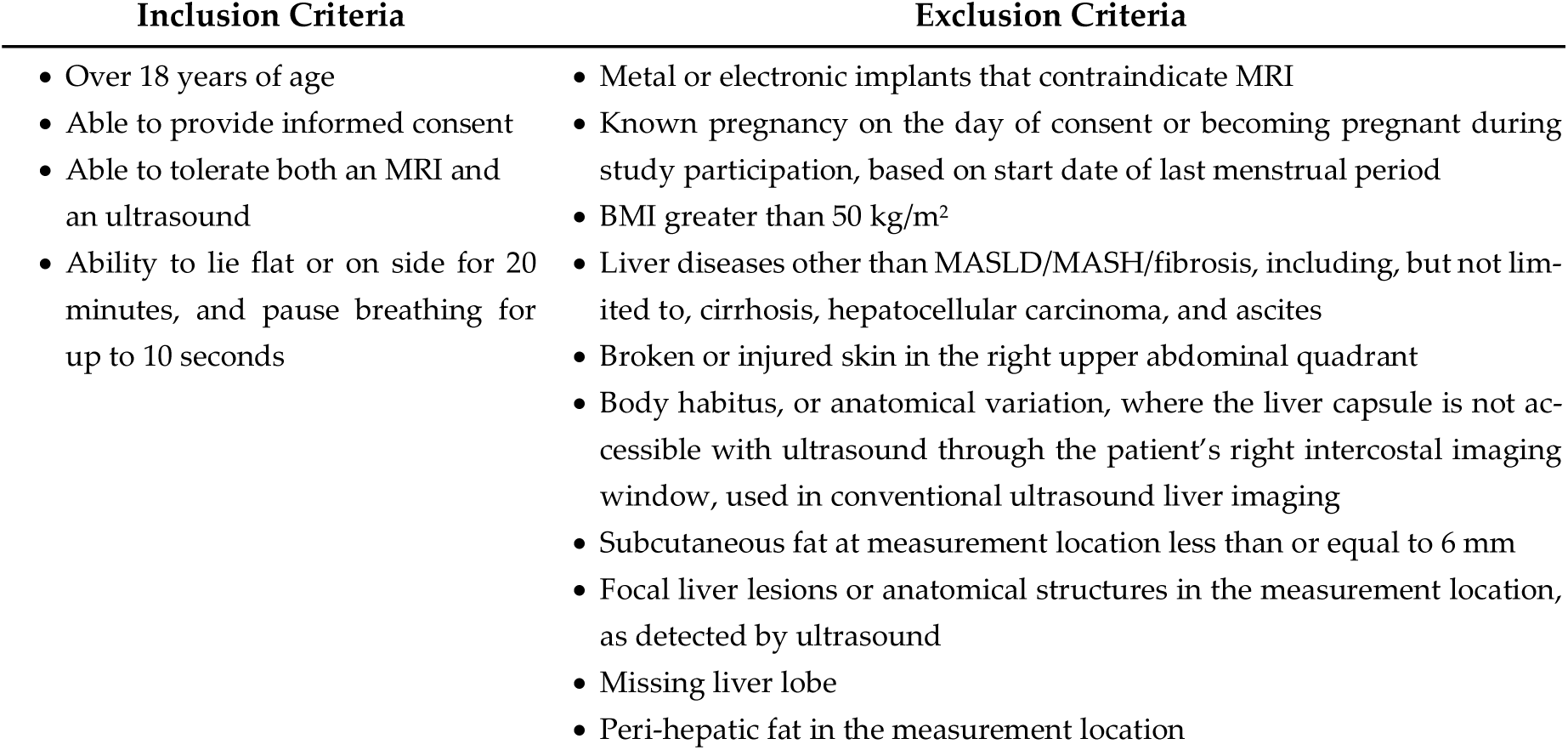
Study inclusion and exclusion criteria.

Subjects with liver diseases other than steatosis, such as cancer or ascites, or abnormalities such as missing liver lobes, polycystic disease (multiple hepatic cysts) were excluded from the study to prevent potential confounding factors. Any subject with an obstruction that prevented imaging of the liver capsule through the right intercostal imaging window was also rejected to ensure adequate measurement of the liver. Subjects with perihepatic fat at the measurement site, as identified by ultrasound or MRI, were also excluded from the study, as the current TA liver fat estimation algorithm assumes no perihepatic fat at that location. Finally, due to the low-frequency transducer (0.5 MHz) of the probe, subjects with subcutaneous fat at the measurement location of less than or equal to 6 mm were also rejected, as this thin fatty layer produced poorly resolved signals at the fat-to-muscle boundary. A flowchart of the study is shown in Figure 2.

**Figure 2.**
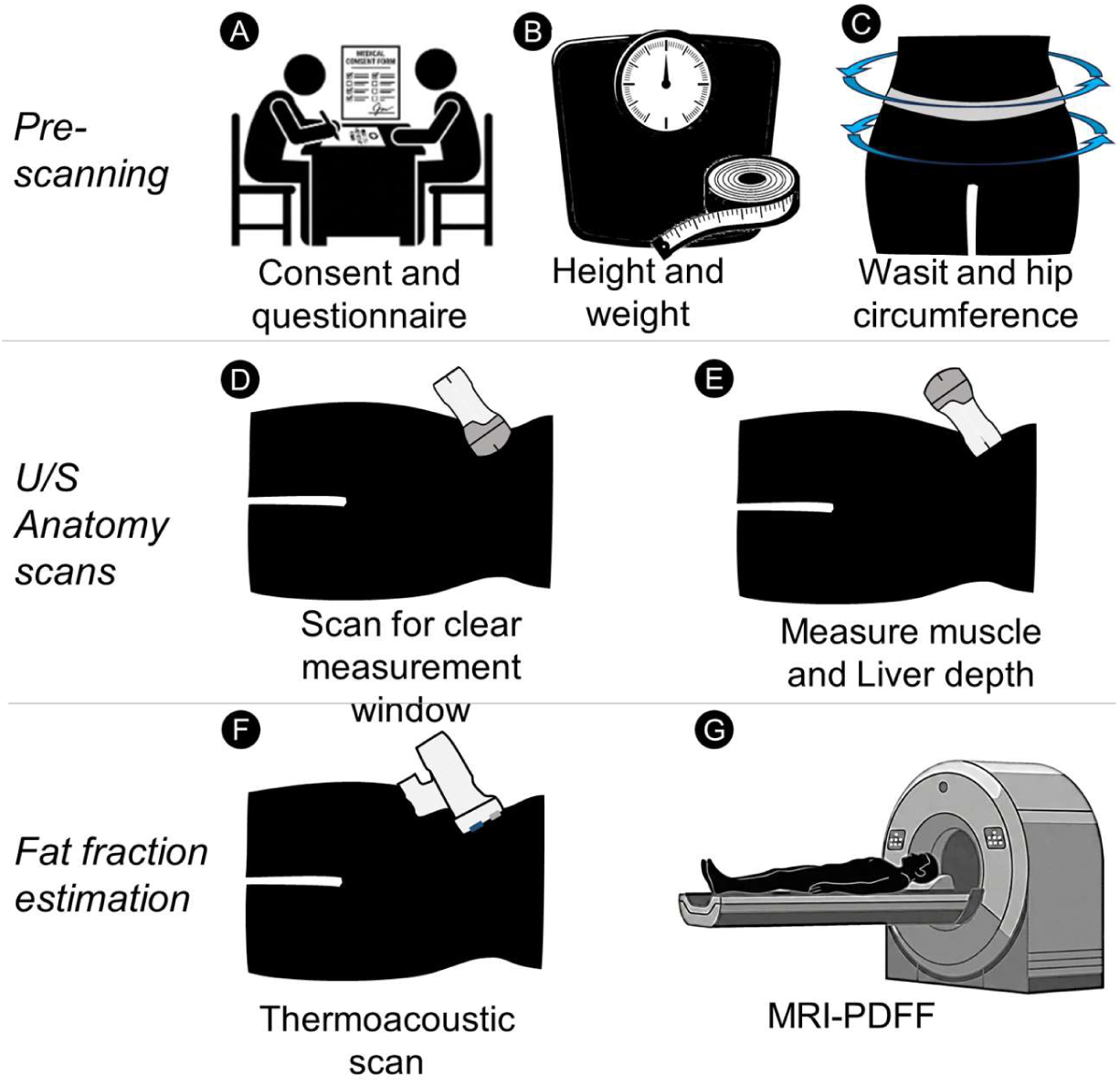
Graphical description of clinical study. (A) Subjects were first consented and their relevant medical history and demographics were recorded (B&C) Body habitus - weight, height, waist, and hip circumference were recorded. (D) A curved array, wireless U/S scanner was used to locate a field of view clear of obstructions such as lung, bowel & perihepatic fat. (E) The wireless, linear array U/S scanner was used to measure the depth of the subcutaneous fat to muscle boundary and the liver capsule boundary location. (F) The probe was then positioned at the exact anatomic location and orientation as the linear array to estimate the liver fat of each subject (G). Each subject was sent to a reference standard MRI-PDFF scan for recording the ground truth fat fraction.

Subjects were instructed to fast for 4 hours prior to the ultrasound scan. The scan was performed by an operator experienced in using the TAEUS system. Subjects were in a slight (30-degree) left lateral decubitus position, with the right arm raised to extend the intercostal space. During a scanning session, the subject’s anatomy was assessed using a wireless, handheld ultrasound system (Vscan Air™ CL, GE Health Care, Milwaukee, WI, USA). A liver window clear of obstructions was first identified with the curved-array side of the probe, after which the subcutaneous fat thickness and liver capsule depth were measured using the linear-array side. The measurement position was marked on the subject with a surgical marker, and the probe orientation was captured with a 9-axis Bluetooth inclinometer (WT901BLECL, WitMotion, Shenzhen, China). Subsequently, a TA scan was performed. A second inclinometer was attached to the TA probe to ensure its orientation matched that of the ultrasound probe. The procedure time for the entire session was usually shorter than 10 minutes.

TAFF and MRI-PDFF evaluations were obtained within a four-week period to minimize changes in the subject’s liver fat. MRI-PDFF images were acquired by certified personnel at the University of Michigan Hospital on a 3.0-T clinical MRI scanner (Ingenia, Philips Healthcare, Netherlands). A three-plane localization gradient-echo sequence was first obtained, followed by an mDIXON-Quant sequence in a single breath hold, which automatically reconstructs the PDFF map. The sequence parameters were: 6 TEs (first TE 0.98 ms, delta TE 0.8 ms), TR 6.3 ms, flip angle 3°, parallel imaging SENSE factor 2, no signal averaging, matrix size 160 × 140, field-of-view (FOV) 400 × 350 mm, 77 slices, and slice thickness 3 mm. MRI-PDFF measurements were obtained from nine manually placed circular regions of interest (ROIs), one in each hepatic segment (I, II, III, IVa, IVb, V, VI, VII, and VIII), ranging from 78 mm² (∼10 mm diameter) to 314 mm² (∼20 mm diameter), while avoiding large blood vessels [60]. The mean PDFF of the nine ROIs was calculated for each scan. University personnel were blinded to the TA LFF results to avoid skewing the comparison.

### Data analysis

Regression, area under the receiver operating characteristic curve (AUROC), and accuracy analyses were conducted to evaluate TAFF’s performance relative to the reference standard MRI-PDFF, with each subject represented by a paired measurement (mean TAFF, mean MRI-PDFF). A Bland-Altman analysis [61] was used to visualize the agreement and distribution of measurement errors. As this clinical study is the first to utilize TAFF, the Bland-Altman plot was also used to identify and correct for measurement bias (i.e., the mean error). Following bias correction, a Deming regression analysis [62] was used to evaluate the relationship between the MRI-PDFF and TAFF estimates of the liver fat. Deming regression was chosen because it appropriately accounts for measurement error in both the reference and test methods, making it well-suited to equivalence testing across methods. Unlike ordinary linear regression, which assumes the independent variable is measured without error, Deming regression provides an unbiased estimate of the relationship between two imperfectly measured variables. It is widely used for comparing clinical and analytical methods and is recommended in several methodological guidelines [63].

To estimate the Deming regression line, the ratio of variances between TAFF and MRI-PDFF measurements was required. Measurement variability was calculated from repeated measurements on multiple subjects. To avoid underestimating the variance of TAFF measurements, subjects with only one valid scan were excluded from the average estimate (35 subjects). As MRI performance can vary across vendors and sites, a conservative approach was adopted, relying on the Radiological Society of North America - Quantitative Imaging Biomarkers Alliance (RSNA-QIBA) publication, which quantified MRI-PDFF’s variance using phantom data [15,64]. These publications suggest a standard deviation of 1.67% for pt fat fraction, corresponding to a variance of 2.778. For living subjects, this variance is expected to increase due to inherent variability among in vivo measurements; thus, the performance reported here represents a lower bound. For the TA, the single-operator repeated-measurement standard deviation was computed within subjects with multiple valid scans and then averaged across subjects. The resulting Deming regression confidence interval was compared with the identity line (MRI-PDFF = TAFF) to assess method equivalence.

In addition, the accuracy of TA in differentiating grades of steatotic liver disease was evaluated using paired MRI-PDFF measurements to estimate sensitivity, specificity, and positive and negative predictive values for binary classification. As there are no widely accepted cut-offs for MASLD disease staging other than S0-S1, cut-offs were selected based on the published studies comparing biopsy staging of SLD to MRI-PDFF measurements, with multiple cutoffs presented based on different publications [65–69]. The steatosis grade cutoffs analyzed are: S0-S1 = 5% LFF, S1-S2 = 12% & 17% LFF, and S2-S3 = 20% & 22% LFF. Additionally, the accuracy of TA in determining whether a subject has ≤8% MRI-PDFF was estimated, as clinical guidelines [70] suggest this MRI-PDFF threshold as one of the decision criteria for therapeutic intervention with Resmetirom. Receiver operating characteristic (ROC) analysis was also performed at all thresholds using Harrell’s C-statistic [71].

Finally, an exploratory analysis was conducted to identify demographic characteristics commonly associated with MASLD that might contribute to measurement error. To determine anatomical and population-based systematic errors, the correlation between measurement error and gender, BMI, and hip-to-waist ratio was assessed.

### Measurement reliability analysis

To characterize system stability, measurement repeatability (intra-operator) and reproducibility (inter-operator) were evaluated in a dedicated study under the IRB committee of Rocky Vista University (IRB approval number: 2018-0029). Two operators (one experienced with the TAEUS system and one a radiologist experienced in U/S imaging). For each subject, two operators independently performed two examinations, with the sequence of operators randomized for each subject to prevent systematic bias. Each of the four examinations per subject was conducted as a completely independent event. Operators were required to fully reset the clinical workflow for every acquisition, which involved repositioning the subject, performing independent ultrasound anatomical localization to identify a clear liver window, identifying distances to tissue boundaries, and finally conducting the thermoacoustic exam. To ensure consistency across the study, a measurement was standardized to require five valid thermoacoustic scans per examination. Scan validity was determined using the scan-quality metric described in Section 2, ensuring that each scan met the necessary geometric and signal-based criteria for accurate TAFF estimation. A single TAFF measurement was defined for analysis as the mean of these five liver fat estimates.

A total of 56 TAFF measurements (14 subjects, 2 operators, 2 repetitions) were included in the final analysis. Reliability and precision were evaluated using a two-way random-effects ANOVA model, which partitioned total variance into components attributable to subjects, operators, and residual error. This model was used to calculate the ICC(2,1) for single measurements. The ICC(2,1) was selected to assess absolute agreement between measurements, ensuring that the results are generalizable to any trained operator in a clinical setting. The Standard Error of Measurement (SEM) was also calculated as the square root of the residual mean square error from the ANOVA model to quantify the precision of an individual measurement. Finally, a Bland-Altman analysis was performed to visualize inter-operator reproducibility, enabling assessment of systematic bias and calculation of 95% limits of agreement across users.

## 3. Results

### Study subjects

Forty-nine (49) subjects consented, enrolled, and were scanned, with eight (8) excluded due to the exclusion criteria shown in Table 1. A preliminary U/S scan determined that three subjects had subcutaneous fat thickness of less than 6 mm. Due to anatomical variation, the liver could not be identified in three excluded subjects who lacked the required right intercostal ultrasound access. Two subjects were excluded due to the presence of perihepatic fat at the measurement site, which was later confirmed by MRI in each case. Finally, one additional subject could not be analyzed due to MRI-PDFF motion artifacts, preventing accurate measurement of LFF. No subject reported a prior diagnosis of MASH or Fibrosis.

The study population is described in Table 2 below. The study population had a higher mean BMI than the US general population, as only subjects with suspected elevated liver fat were enrolled. The mean BMI of the subjects was 34 kg/m² in females and 32.2 kg/m² in males, compared with the national average of 29.6 kg/m² [72]. Two-thirds of the subjects are obese (BMI > 30). Waist circumference was also larger than the national average and exceeded the threshold for abdominal obesity. For females, the mean waist circumference was 113 cm, substantially exceeding the national average of 95.5 cm and the cutoff for abdominal obesity of 88 cm. For males, the mean waist circumference was 112 cm or higher, exceeding the national average of 100.5 cm and the cutoff for abdominal obesity of 102 cm [72].

**Table 2.** Demographics and Body Habitus of Subjects.

| Gender | N | Variable | Units | Mean | Std Dev | Minimum | Maximum |
| --- | --- | --- | --- | --- | --- | --- | --- |
| Female | 25 | Age | years | 54.2 | 12.0 | 34.0 | 74.0 |
|  |  | Height | cm | 165.7 | 7.6 | 153.0 | 184.0 |
|  |  | Weight | kg | 93.6 | 22.0 | 54.0 | 148.8 |
|  |  | BMI | kg/m2 | 34.0 | 7.1 | 21.0 | 47.0 |
|  |  | Hip circumference | cm | 120.3 | 16.7 | 84.0 | 150.0 |
|  |  | Waist circumference | cm | 113.4 | 16.8 | 77.0 | 148.0 |
|  |  | Waist to hip ratio | # | 0.95 | 0.09 | 0.79 | 1.17 |
| Male | 15 | Age | years | 49.2 | 10.3 | 29.0 | 65.0 |
|  |  | Height | cm | 176.5 | 8.0 | 157.0 | 186.0 |
|  |  | Weight | kg | 100.5 | 14.3 | 79.2 | 130.0 |
|  |  | BMI | kg/m2 | 32.2 | 3.7 | 24.8 | 37.6 |
|  |  | Hip circumference | cm | 112.0 | 6.1 | 103.0 | 123.0 |
|  |  | Waist circumference | cm | 112.1 | 13.8 | 83.0 | 134.0 |
|  |  | Waist to hip ratio | # | 1.01 | 0.09 | 0.81 | 1.13 |
| Both | 40 | Age | years | 52.3 | 11.5 | 29.0 | 74.0 |
|  |  | Height | cm | 169.8 | 9.3 | 153.0 | 186.0 |
|  |  | Weight | kg | 96.2 | 19.6 | 54.0 | 148.8 |
|  |  | BMI | kg/m2 | 33.3 | 6.0 | 21.0 | 47.0 |
|  |  | Hip circumference | cm | 117.1 | 14.2 | 84.0 | 150.0 |
|  |  | Waist circumference | cm | 112.9 | 15 | 77.0 | 148.0 |
|  |  | Waist to hip ratio | # | 0.97 | 0.09 | 0.79 | 1.17 |

Due to enrollment criteria, the incidence of MASLD in this study was higher than that of the United States population [15]. The distribution of liver steatosis grades is shown in Figure 3(A). This more uniform distribution of MASLD grades enabled adequate estimation of TA performance across all MASLD grades. Similarly, the distributions of body habitus (subcutaneous fat thickness and liver capsule depth) shown in Figures 3(B) and 3(C), respectively, indicate a greater tendency toward a larger body habitus.

**Figure 3.**
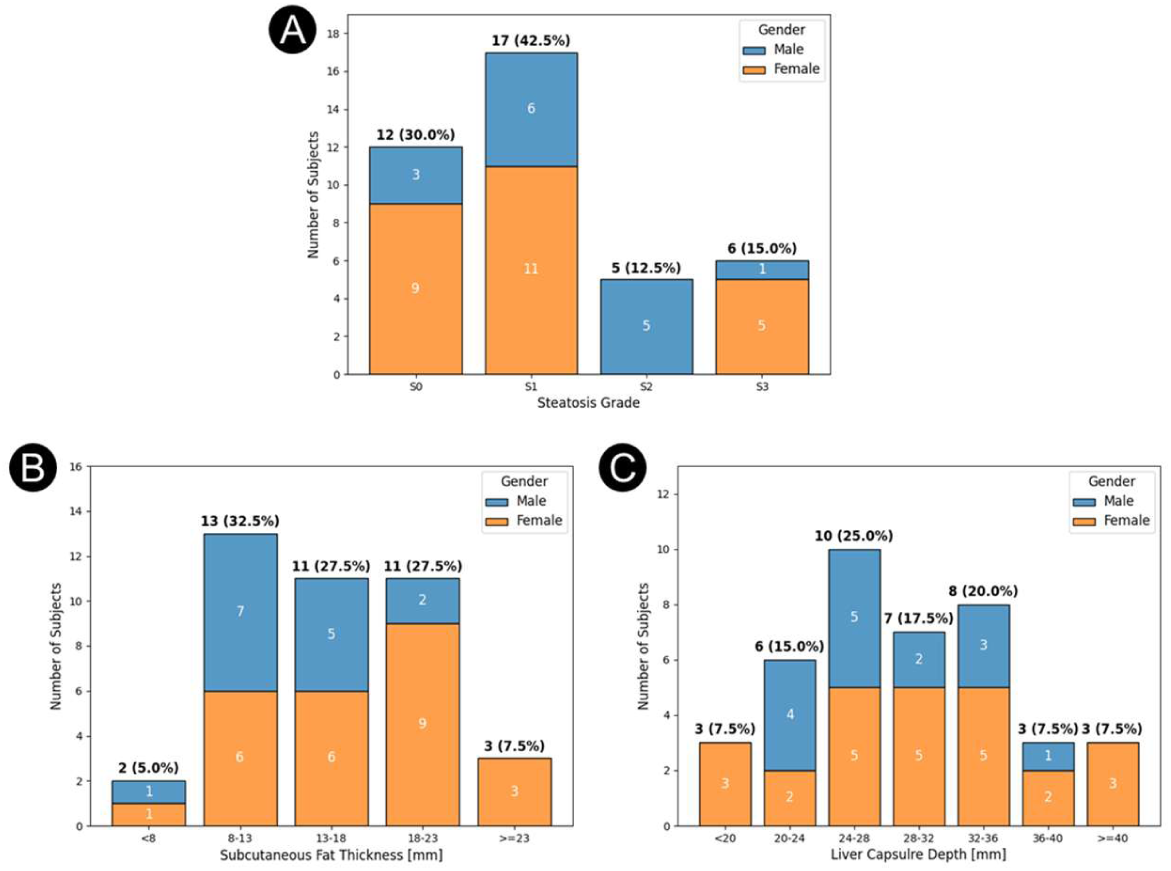
Study population characteristics by gender: (A) Steatosis grade (by MRI-PDFF) of the subjects, (B) Subcutaneous fat thickness (by U/S), (C) Liver capsule depth (by U/S)

### TAFF performance

The mean estimated TAFF value and the paired MRI-PDFF measurement were used to generate a Bland-Altman plot to visualize the distribution of error across MASLD grades. The mean TAFF measurement error for the 40 subjects who met the inclusion criteria was 1.95% LFF. This 1.95% error was subtracted from each subject’s TAFF value to correct for bias. This systematic offset is attributed to the transition from theoretical models and idealized simulations to actual in vivo human data. To further characterize measurement variability and investigate potential dependencies, a separate Repeatability and Reproducibility (R&R) study was conducted; details of which are provided in the following section. Figure 4A shows that across a wide range of LFF (1%-32%), and for all steatosis grades (colored differently), the average absolute error is 3.04%. TAFF measurement error was higher in subjects with low MRI-PDFF values than in those with higher LFF. The limits of agreement (LOA) were ±7.21% LFF. Figure 4B illustrates the distribution of absolute error across LFF. It represents the percentage of subjects whose measurement error falls below a given threshold. Percentiles were calculated via linear interpolation of the empirical cumulative distribution function (folded CDF). From the plot, we observe that 49.1% of subjects had a TAFF estimate within 2.5% of their MRI-PDFF value, and 89.1% within 5.0%. These plots illustrate the accuracy and consistency of the TAFF estimates of LFF across the study population compared with MRI-PDFF measurements.

**Figure 4.**
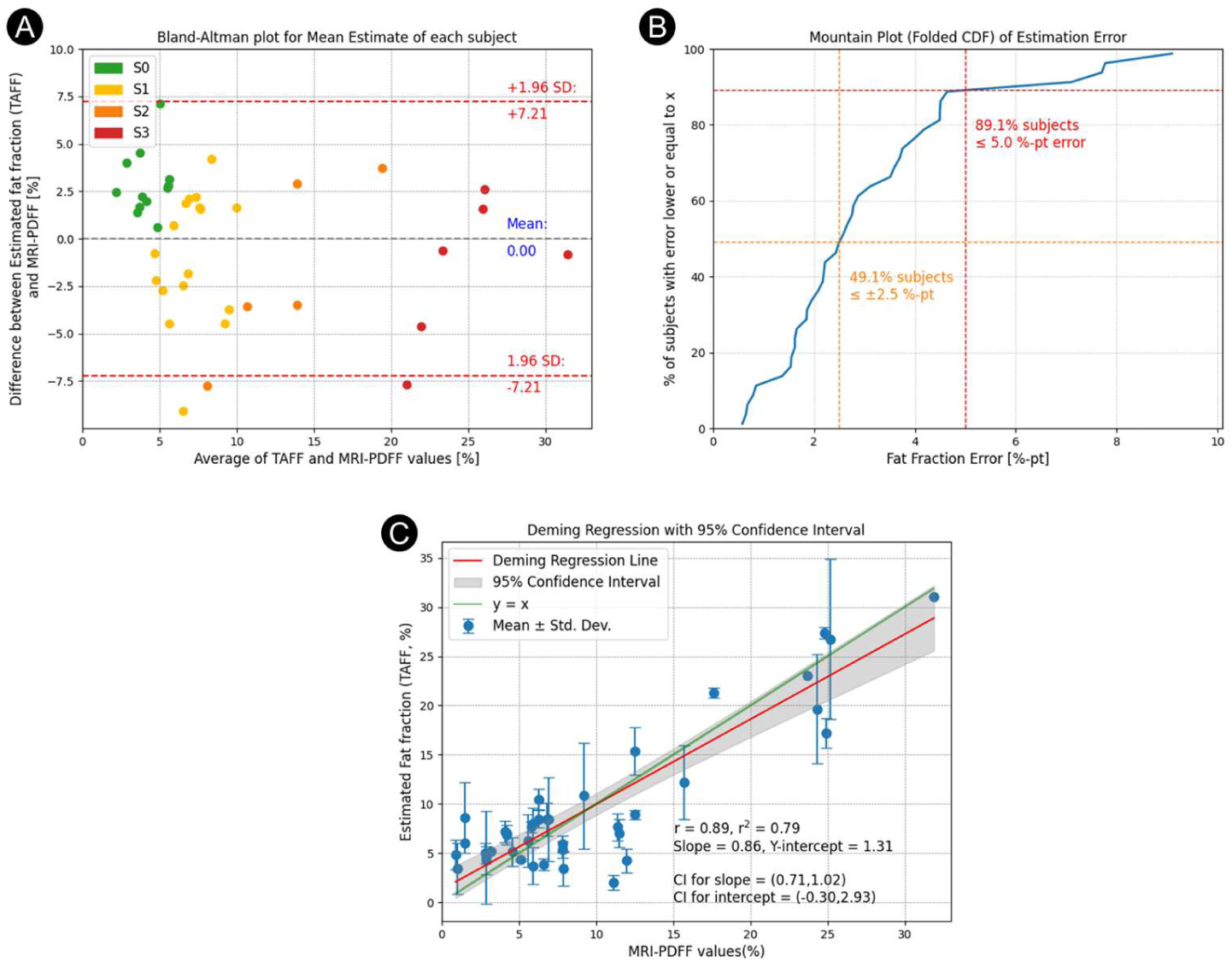
Analysis of study results in comparison to MRI-PDFF. (A) Bland-Altman plot of the estimation error (in % fat fraction) as a function of the averaged fat fraction. (B) Error distribution plot (C) Deming regression plot

The within-subject variability of TAFF was assessed, yielding an average standard deviation of 2.66% (variance of 7.085) LFF for repeated measurements with TA for subjects with more than a single valid scan. Deming regression, accounting for TAFF’s higher variability relative to MRI-PDFF (standard variation of 1.667% or variance of 2.778, yielding a TAFF to MRI-PDFF variance ratio of 2.55), was used to evaluate agreement between TAFF and MRI-PDFF estimates of LFF, as shown in Figure 4B. The Deming regression yielded a slope of 0.86 (95% confidence interval: 0.71 to 1.02) and an intercept of 1.31% LFF (95% confidence interval: -0.30 to 2.93), which includes the identity line, indicating good agreement between TAEUS and MRI-PDFF. In addition, the Pearson correlation coefficient was determined to be 0.89, further demonstrating a strong correlation between TAFF and MRI-PDFF measurements of LFF. The within-subject variability of repeated TAFF estimates was higher at low LFF than at higher LFF.

The classification accuracy of TAFF across different LFF thresholds is summarized in Table 3 below. For each value, the 95% confidence interval was also calculated and presented in parentheses. Consistent with the earlier observation that signal variability is higher at low fat fractions, classification performance is limited at the lowest LFF threshold (5%). As LFF increases, both sensitivity and specificity improve, resulting in higher overall accuracy and a larger AUROC. The positive predictive values are good across the board, and the negative predictive values increase markedly at moderate fat fractions. Furthermore, as illustrated in Figure 5, the ROC curves show a marked improvement at higher fat fraction thresholds, accompanied by corresponding increases in AUROC, reflecting enhanced classification performance of TAFF for moderate-to-severe liver fat.

**Figure 5.**
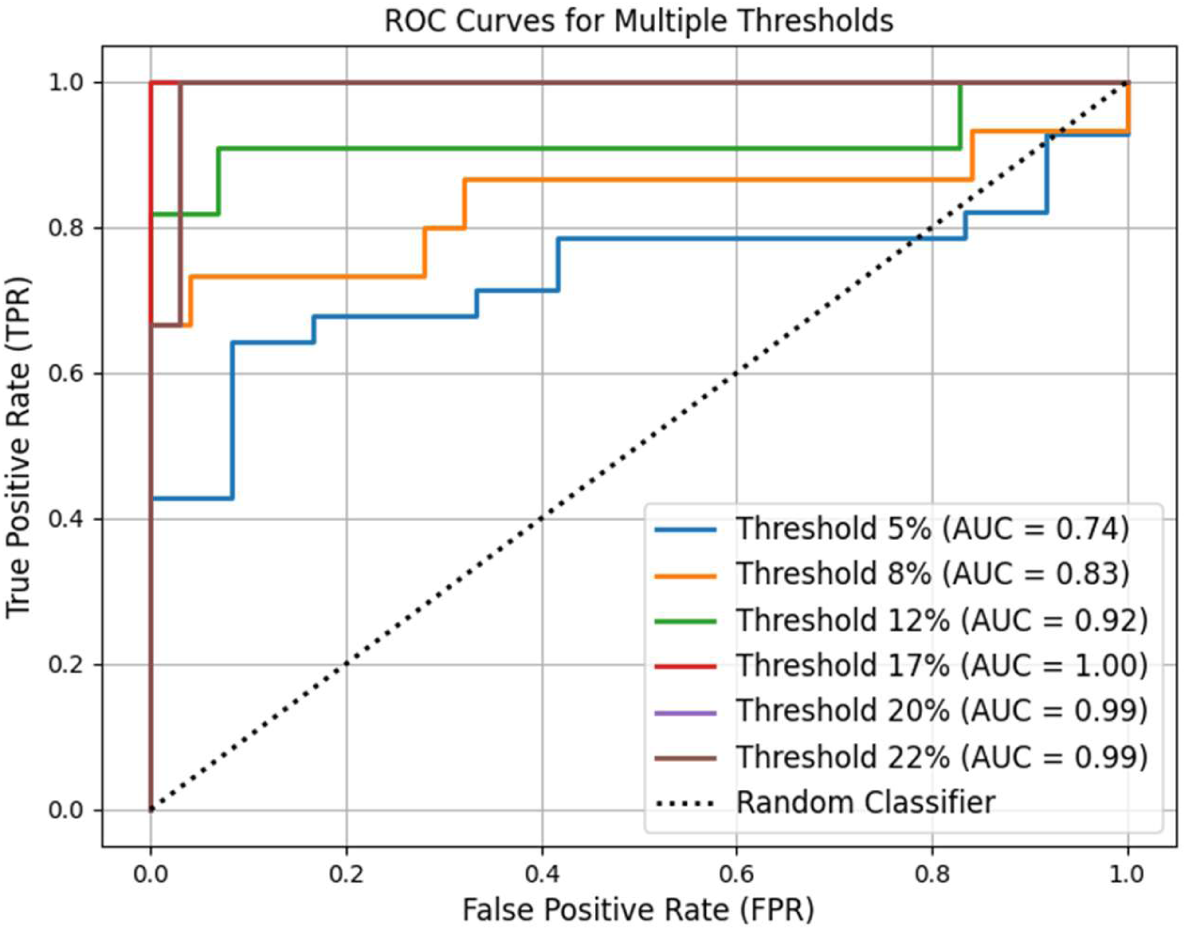
ROC curves for the binary classification results (Thresholds: = 5%, 8%, 12%, 17%, 20%, and 22%)

**Table 3.** Binary classification results with various threshold values.

| Threshold | 5% LFF:<br>S0-S1 | 8% LFF:<br>THrβ | 12% LFF:<br>S1-S2 | 17% LFF:<br>S1-S2 | 20% LFF:<br>S2-S3 | 22% LFF:<br>S2-S3 |
| --- | --- | --- | --- | --- | --- | --- |
| PPV [%] | 75.9<br>(59.3, 90.6) | 68.8<br>(43.8, 90.9) | 100.0<br>(100.0, 100.0) | 100.0<br>(100.0, 100.0) | 80.0<br>(33.3, 100.0) | 100.0<br>(100.0, 100.0) |
| NPV [%] | 45.5<br>(16.7, 75.0) | 83.3<br>(66.7, 96.3) | 93.5<br>(83.3, 100.0) | 100.0<br>(100.0, 100.0) | 94.3<br>(85.7, 100.0) | 94.4<br>(86.1, 100.0) |
| **Sensitivity [%]** | 78.6<br>(62.1, 92.6) | 73.3<br>(50.0, 94.1) | 81.8<br>(54.55, 100.0) | 100.0<br>(100.0, 100.0) | 66.7<br>(20.0, 100.0) | 66.7<br>(20.0, 100.0) |
| **Specificity [%]** | 41.7<br>(12.5, 72.7) | 80.0<br>(63.0, 95.0) | 100.0<br>(100.0, 100.0) | 100.0<br>(100.0, 100.0) | 97.1<br>(90.3, 100.0) | 100.0<br>(100.0, 100.0) |
| **Accuracy [%]** | 67.5<br>(52.5, 82.5) | 77.5<br>(65.0, 90.0) | 95.0<br>(87.5, 100.0) | 100.0<br>(100.0, 100.0) | 92.5<br>(82.50, 100.0) | 95.0<br>(87.5, 100.0) |
| **AUROC [#]** | 0.74<br>(0.57, 0.88) | 0.83<br>(0.66, 0.98) | 0.92<br>(0.75, 1.00) | 1.0<br>(1.0,1.0) | 0.99<br>(0.96, 1.00) | 0.99<br>(0.96, 1.00) |

### Relationship between TAFF and demographic information

Differences between TAFF and MRI were analyzed with respect to demographics. We found that the average difference in males (n = 15) was 1.6% LFF lower than in females (n = 25), yielding a p-value of 0.26. This finding suggests that there is no statistically significant difference between the genders. Additionally, a statistical analysis was conducted to investigate the relationship between the various metadata collected on subjects and the estimation differences as reported in Table 4 below.

**Table 4.**
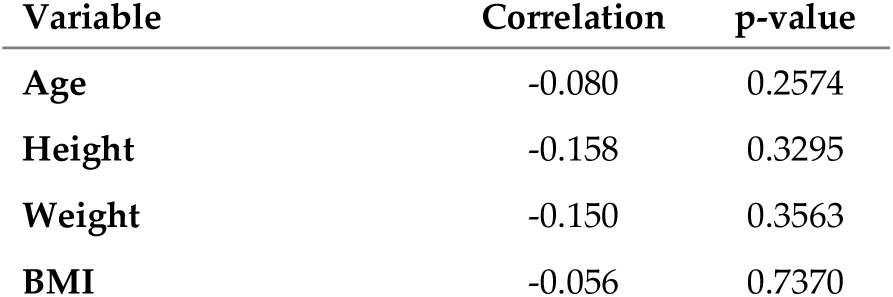
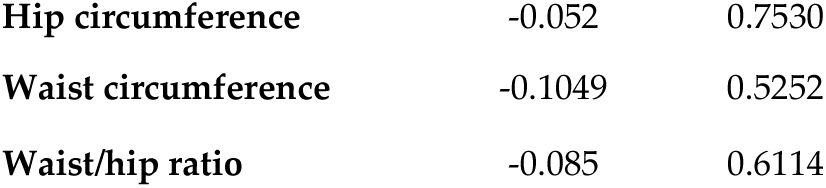
Relationship between meta-variables and estimation errors.

| Variable | Correlation | p-value |
| --- | --- | --- |
| Age | -0.080 | 0.2574 |
| Height | -0.158 | 0.3295 |
| Weight | -0.150 | 0.3563 |
| BMI | -0.056 | 0.7370 |
| Hip circumference | -0.052 | 0.7530 |
| Waist circumference | -0.1049 | 0.5252 |
| Waist/hip ratio | -0.085 | 0.6114 |

This analysis found low correlations and no statistically significant relationships between metadata variables and TA estimation errors. Figure 6 presents the distribution of TAFF estimation errors, categorized by subjects’ demographic and anthropometric characteristics. These box plots illustrate the median, interquartile range, and outliers for each sub-group (gender, height, weight, BMI, and body circumference metrics). Consistent with the low correlation coefficients (r < 0.16) and non-significant p-values (p > 0.25) previously noted, the plots show no systematic trends or distinct separation between categories. While specific subgroups, such as the 90 to 110 kg weight class or the 30 to 35 BMI range, exhibit greater variability, the substantial overlap in error distributions across those bins reinforces the conclusion that these metadata variables are unreliable predictors of TAFF estimation performance.

**Figure 6.**
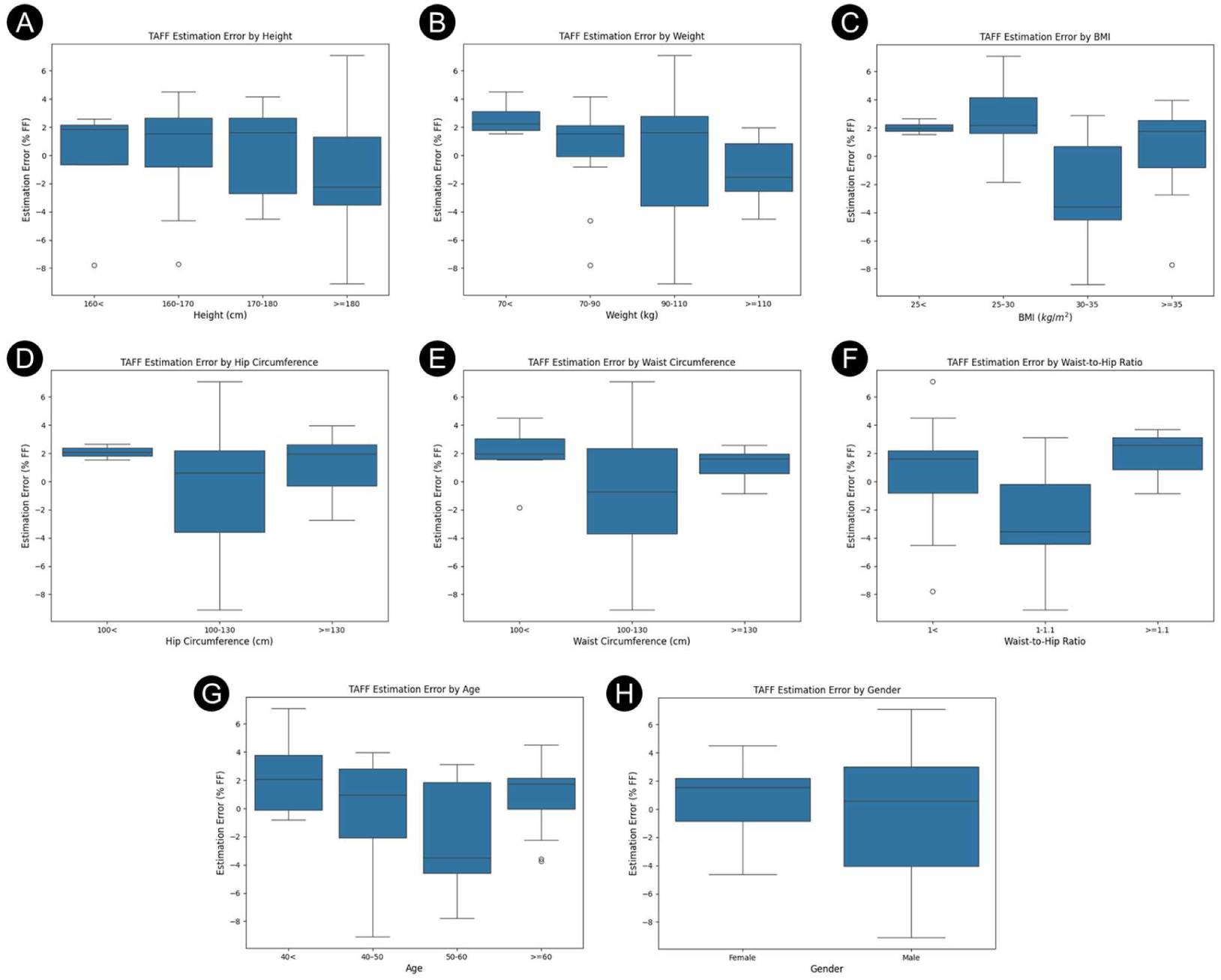
Box Plots of TAFF Estimation Errors Across Demographic and Anthropometric subgroups (A) Height (B) Weight (C) BMI (D) Hip circumference (E) Waist circumference (F) Waist-to-Hip ratio (G) Age (H) Gender

### Measurement reliability

The reliability of the TAFF estimation was evaluated on a total of 56 measurements obtained from 14 subjects. The two-way random-effects ANOVA model yielded an ICC(2,1) of 0.89 (95% CI: 0.74–0.93), indicating good reliability for clinical liver fat quantification.

The system’s precision was characterized by a SEM of 3.31%, similar in magnitude to the 3.04% average absolute error observed in the primary feasibility cohort, suggesting comparable overall measurement variability across datasets.

Interoperator reproducibility was further visualized via Bland-Altman analysis (Figure 7). The analysis revealed high agreement between operators, with a mean difference of only 0.36% and 95% limits of agreement ranging from -5.82% to 5.09%. The mean absolute error between operators was 2.28%. These results demonstrate that the TAEUS system provides stable and consistent estimates across different users and independent scan set-ups.

**Figure 7.**
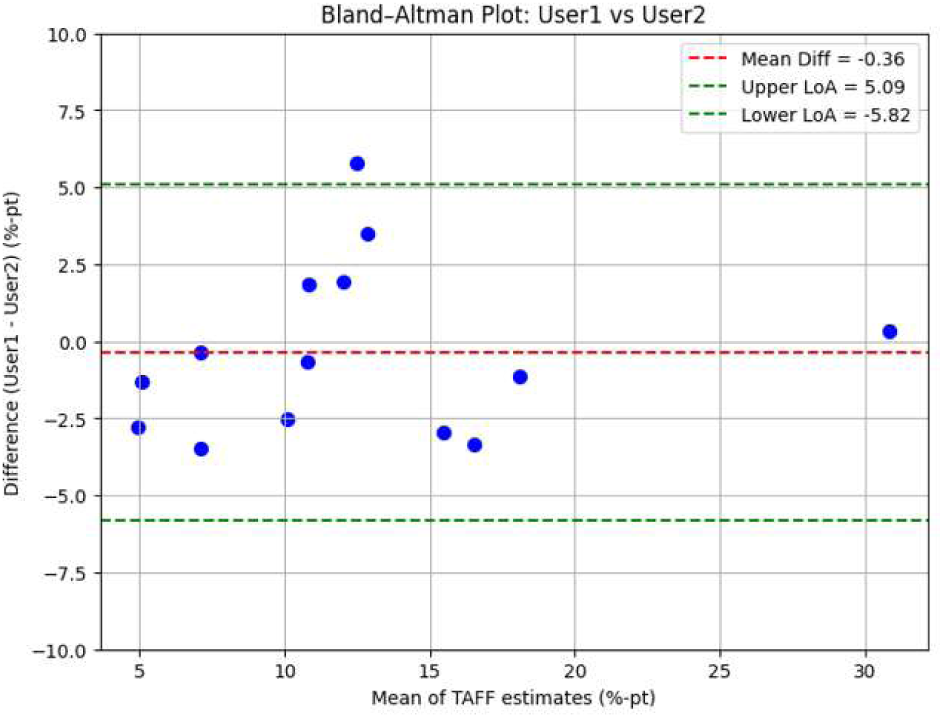
Bland-Altman plot of inter-operator reproducibility (N = 14). The red dashed line indicates a mean difference of 0.36%. Green dashed lines represent the 95% limits of agreement from -5.82% to 5.09%.

## 4. Discussion

This single-site feasibility study is the first of its kind to utilize thermoacoustic principles to estimate fat in human subjects. The TA liver system accurately estimates liver fat across a wide range of body habitus, including mild to severe disease. The portable TA liver system provides liver fat estimates that are consistent with MRI-PDFF, with costs and mobility similar to those of cart-based ultrasound. Thus, it indicates the potential of such technology as a valuable tool to aid in the diagnosis of MASLD, to inform therapeutic decisions, and to monitor disease progression.

Bland-Altman plots across all disease grades show the limits of agreement to be ±7.21% as compared to the reference standard MRI-PDFF. The cumulative error distribution (mountain plot) indicates that half (49.1%) of subjects had a TAFF estimate within 2.5% of their MRI-PDFF value, and the vast majority (89.1%) within 5.0%. Results demonstrate narrower limits of agreement than those reported for CAP, which exhibits a 95% limit of agreement for ±12% in fat fraction after converting its native measurements to match the MRI [73]. More importantly, the TA estimates of LFF were strongly correlated with MRI-PDFF scores (r = 0.89). In contrast, quantitative ultrasound–based estimates of LFF have shown substantial variability, with reported correlations with MRI-PDFF ranging from 0.49 to 0.89 at baseline [74–77], and more limited correlation with longitudinal changes in MRI-PDFF (r ≈ 0.52) [74,75]. In contrast to these quantitative ultrasound approaches, TAFF estimates in the present study exhibit approximately constant bias and do not demonstrate evidence of proportional bias, as indicated by the Bland–Altman analysis.

The 1.95% systematic bias identified via Bland-Altman analysis reflects the transition from idealized theoretical models to actual in vivo human data. While the specific contributions of device, operator, or cohort factors remain to be characterized, data from the separate Repeatability and Reproducibility (R&R) study provide evidence of robust measurement stability. The analysis yielded an ICC(2,1) of 0.89 (95% CI: 0.74–0.93) and a SEM of 3.31%. Notably, this SEM closely aligns with the 3.04% average absolute error observed in the primary cohort, suggesting that measurement precision is a fundamental characteristic of the current system configuration rather than a result of operator technique. In future multi-center studies, these data-driven refinements will be integrated into automated calibration protocols to ensure consistent performance across diverse populations without the need for manual bias correction.

In predicting the presence of moderate to severe steatosis (S2-S3) at the 12% LFF threshold, TAFF scores achieve 100% specificity and PPV of 100%, with an AUROC of 0.92 in the available cohort. Although derived from a limited sample, these results indicate exceptional accuracy in identifying subjects with moderate-to-severe SLD, a critical clinical threshold for urgent and aggressive management of MASLD and MASH. Although this study did not include a direct ‘head-to-head’ comparison with QUS, literature shows, through a large meta-analysis of widely used point-of-care quantitative U/S methods for assessing moderate steatosis, reports AUROCs of approximately 0.82 and lower specificities ranging from 74% to 79% [78,79]. While these existing tools have a role in screening, their relatively low specificity often necessitates confirmatory testing, in contrast with TA LFF estimation. The data gathered in this study suggests that the TAFF has the potential to offer high diagnostic accuracy for quantifying moderate liver fat (AUROC >0.90) as it has a similar contrast mechanism as MRI-PDFF. Thus, it might overcome some of the limitations of current quantitative U/S and provide superior performance.

Performance in the low liver fat range is inherently more variable because the diminished contrast between lean liver and muscle results in a lower contrast-to-noise ratio. In this feasibility assessment, the system achieved an accuracy of 77.5% and an AUROC of 0.83 at an 8% MRI-PDFF threshold. While these results indicate that detection in the ’lean’ population remains challenging with the current system configuration, the negative predictive value (NPV) of 83.3% at this threshold provides preliminary evidence of the technology’s potential to rule out significant steatosis. These findings help define the system’s current operating envelope, demonstrating that while the methodology shows promise for identifying moderate-to-severe steatosis, additional refinements are necessary to improve precision and reliability for early-stage MASLD assessment.

Demographic analysis revealed no significant difference in errors between male and female subjects. In addition, the TAFF scores demonstrated low correlation (r<0.16) between estimation errors and subjects’ age or body habitus parameters (e.g., height, BMI, waist circumference). While a larger body habitus and increased liver capsule depth (as shown in Figure 3) can theoretically reduce the signal-to-noise ratio by attenuating RF energy, these factors did not significantly impact estimation accuracy in this cohort. This implies that the method is applicable across various body types, age groups, and sexes. For QUS, reported clinical findings indicate a significant dependence of LFF measurement accuracy on BMI [77,80], which was not demonstrated by the TAEUS liver system in this study. UGAP, for example, demonstrates markedly lower accuracy for diagnosing severe steatosis (S3) in subjects with BMI > 30 kg/m² [76]. These BMI-associated limitations arise from increased skin-to-capsule distance that degrades acoustic signal quality [12], depth-dependent variability in measurement thresholds, and anthropometric factors that independently affect whether CAP measurements achieve acceptable interquartile ranges [81]. Additional BMI-related tissue characteristics further impair liver fat quantification for these techniques: Karlas et al. [82] described “muscular viscoelastic adaptations” in overweight individuals that alter acoustic propagation, and Maar et al. showed that hepatic imaging quality decreases with increasing BMI. However, high-performance probes can partially mitigate this effect [83]. Consistent with these findings, Cusi et al. [84] reported that ultrasound exhibits low sensitivity for detecting hepatic steatosis in obese and severely obese patients. Depth dependence compounds these challenges, as a retrospective study of acoustic attenuation coefficient algorithms from Canon, Philips, and Siemens demonstrated a progressive decline in AC values with increasing measurement depth, as well as significantly higher values with a 1 cm ROI compared with a 3 cm ROI [85]. In addition, some quantitative ultrasound methods showed a nonlinear relationship (i.e., proportional error) with MRI-PDFF [86,87]. Collectively, these BMI- and depth-related constraints highlight the need for standardized acquisition protocols and improved quantitative ultrasound approaches capable of producing accurate and reliable liver fat measurements across diverse body habitus. Unlike all other imaging techniques except perhaps MRI, thermoacoustics is strictly a fat-specific parameter that does not rely on non-fat-related tissue properties to produce contrast.

These overall findings support TA as a potentially accurate, non-invasive, point-of-care solution that could address the cost and access constraints of MRI and the limitations of current quantitative ultrasound methods. The high PPV and NPV allow for the accurate identification of individuals with severe MASLD and for separating healthy individuals and those with mild MASLD from individuals with moderate and severe MASLD. These trends indicate that TAFF is most effective at detecting moderate to severe steatosis, whereas classification at very low fat fractions remains more challenging due to inherently weaker signals. Specifically, at an LFF of 8%, the Resmetirom (THrβ) treatment threshold, the negative predictive value exceeds 83%, suggesting that TA may be valuable in guiding therapeutic decisions and monitoring disease progression.

The current feasibility study also highlights some limitations of thermoacoustic imaging and the first-generation TA system. In subjects, the current system is not suitable as an early screening tool since it can only resolve contrast differences in tissue conductivity. Thus, it suffers from a lack of signal at low LFF as the differences between lean liver tissue and (lean) muscle become minute, leading to a lower contrast-to-noise ratio and more variable measurements. As liver tissue becomes fattier, the contrast increases, and with it, the robustness of the liver fat estimation. This limitation of the first-generation thermo-acoustic system can be addressed by incorporating a more sensitive transducer to improve signal resolution for low LFF. The current 16-element, 0.5 MHz phased-array configuration offers limited performance that can be improved. In addition, the improvement in contrast with increasing fat fraction, rather than extinguishing signal observed with pure ultrasonic methods [77], suggests that a combined quantitative ultrasound and TA device can deliver improved performance across all steatosis grades.

Another difficulty with this first-generation TA system is the need to rely on a separate U/S device to acquire tissue anatomy before a TA scan can proceed. Proper positioning of the wireless ultrasound probe and the larger TA probe contributed substantially to estimation inaccuracies, lengthened the session time, and imposed additional strain on the user. This observation suggests that a future device combining both ultrasound and TA capabilities would be much easier to use and would achieve greater accuracy in fat fraction estimation. It is unclear if fibrosis has a non-negligible effect on TA measurements. The TA signal amplitude is proportional to the tissue conductivity, which decreases with liver fibrosis [88–90], but also to the tissue bulk modulus (through the Grüneisen parameter), which increases with fibrosis [91,92]. In addition, liver fibrosis and other potential confounders should be thoroughly investigated in future studies to ensure TA liver fat estimation is applicable to a wide range of patients. Fibrosis can affect permittivity, perturbing the amount of electromagnetic power that reaches and is deposited in the liver.

It is also worth noting that while the results reported in this study are quite promising, they are based on a preliminary study with a single site and a limited number of subjects (N = 40). Moreover, the current study excluded subjects with liver fibrosis to minimize this confounder, relying on self-reported medical history. It is worth noting that many patients with steatosis may be unaware of their fibrosis status, which may contribute to inaccuracies in the results. Future studies should include direct measurement of fibrosis to address that. Although the current work successfully established the system’s initial reliability through a dedicated R&R study, demonstrating good agreement (ICC = 0.89) and a negligible mean inter-operator difference of 0.36%, a larger-scale, multi-site, multi-user follow-up study remains essential to further validate these findings across broader clinical environments. In addition, direct comparison with QUS by a qualified sonographer would allow better benchmarking of the different techniques, thus outlining the benefits and drawbacks of each method. Thus, it is expected that in the coming years, more studies will be conducted using TA, opening new avenues for accurate, low-cost, portable liver fat estimation and monitoring.

## Author Contributions

Conceptualization, J.H.C., M.T. and I.S.; methodology, J.H.C., C.M.B., M.T. and I.S.; software, J.H.C.; validation, All Authors.; formal analysis, J.H.C., J.G., and I.S.; investigation, C.M.B, J.G, and J.M.R.; resources, M.T., J.M.R., and J.G.; data curation, J.H.C. and M.T.; writing—original draft preparation, J.H.C., C.M.B., M.T., and I.S.; writing—review and editing, J.M.R. and J.G.; visualization, J.H.C. and I.S.; supervision, I.S.; All authors have read and agreed to the published version of the manuscript.

## Institutional Review Board Statement

The feasibility study was conducted in accordance with the Declaration of Helsinki, and approved by the Advarra Institutional Review Board (IRB approval number: Pro00089043), obtained 08/06/2025 The R&R study was conducted in accordance with the Declaration of Helsinki, and approved by the IRB committee of Rocky Vista University (IRB approval number: 2018-0029), with the latest amendment obtained 01/06/2026.

## Informed Consent Statement

Informed consent was obtained from all subjects involved in the study.

## Data Availability Statement

The original contributions presented in this study are included in the article - Appendix B. Further inquiries can be directed to the corresponding author.

## Acknowledgments

The authors express their gratitude to Chris Davis, PhD, for his review and meaningful improvements to this manuscript, and to Professor Emeritus Gary Cutter for his statistical review of the data analysis. We also thank Professor Thomas L. Chenevert of the Department of Radiology at the University of Michigan for his technical assistance in acquiring MRI-PDFF measurements. Finally, we acknowledge Ms. Ava Ohlgren for her invaluable support in recruiting, scheduling, and accompanying the study subjects.

## Conflicts of Interest

The authors declare no conflicts of interest. J.H.C., M.T., and I.S. were, at the time of research, employees of Endra Life Sciences, Inc. C.M.B. was, at the time of the research, an independent consultant contracted by Endra Life Sciences, Inc. J.M.R. and J.G. were, at the time of research, paid Scientific advisors for Endra Life Sciences.

The following abbreviations are used in this manuscript:

## Abbreviations

AASLD: American Association for the Study of Liver Diseases
AC: Attenuation Coefficient
BMI: Body Mass Index
CAP: Controlled Attenuation Parameter
CDF: Cumulative Distribution Function
CVD: Cardio-Vascular Disease
EM: Electro-Magnetic
GLP-1: RA Glucagon-Like Peptide-1 Receptor Agonists
HCC: Hepato-Cellular Carcinoma
ICC: Intraclass Correlation Coefficient
LFF: Liver Fat Fraction
LOA: Limits Of Agreement
MASH: Metabolic Dysfunction-Associated Steatohepatitis
MRI-PDFF: Magnetic Resonance Imaging-Proton Density Fat Fraction
MASLD: Metabolic Dysfunction-Associated Steatotic Liver Disease
MWA: Micro-Wave Ablation
NAFLD: Non-Alcoholic Fatty Liver Disease
NPV: Negative Predictive Value
QIBA: Quantitative Imaging Biomarkers Alliance
QUS: Quantitative Ultra-Sound
RF: Radio-Frequency
ROI: Region of Interest
RSNA: Radiological Society of North America
R&R: Repeatability and Reproducibility
SEM: Standard Error of Measurement
TA: Thermo-Acoustic
TAEUS: Thermo-Acoustic Enhanced Ultrasound System
TAFF: Thermo-Acoustic Fat Fraction
T2D: Type 2 Diabetes
US: United States
U/S: Ultra-Sound
UDFF: Ultrasound-Derived Fat Fraction
UGAP: Ultrasound-Guided Attenuation Parameter

## Appendix A

### Detailed description of TAFF estimation

The thermoacoustic signal amplitude P generated from a tissue can be modeled as follows:

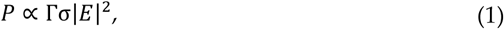

Where 𝛤 and σ are the Gruneisen parameter and the conductivity of a given tissue, respectively, and |E| is the electric field strength for the tissue. Both tissue properties and electric field strengths depend on spatial location. For simplicity, the dependency on the location will be omitted.

The TAEUS probe has a limited field-of-view angle and a narrow-band ultrasound transducer. As a result, the TAEUS system is only sensitive to changes in tissue properties and to signals generated at tissue boundaries. The thermoacoustic signal at a tissue boundary can be modeled as a difference between signals generated at each tissue:

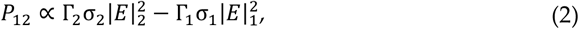

Where subscripts 1 and 2 denote tissues on each side of the boundary.

To estimate the fat fraction of the liver of a subject, we need to collect a signal from the boundary between the liver and a different tissue. We position the TAEUS probe on the subject’s body so that the field of view includes a tissue boundary between the muscle and liver layers. The signal at this boundary can be modeled based on equation (2) as follows:

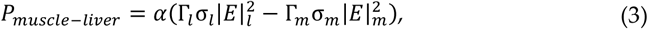

where α is a system-dependent coefficient and subscripts “m” and “l” denote muscle and liver, respectively. Since the electric field within the body cannot be measured, it must be estimated. Using Ansys, we modeled various body habitus with varying degrees of liver fat to estimate the electric field strength distribution of a given subject. To compensate for simplified modeling and various assumptions made, we also considered a natural reference point within the body. Instead of estimating liver fat using the above equation, another boundary based on known tissue properties was considered. The subcutaneous fat-to-muscle boundary always exists in the selected field of view containing the muscle-to-liver boundary. Since the tissue properties of both subcutaneous fat and muscle are known, the signal at this boundary can be used for calibration. We consider the ratio of signals from these boundaries as follows:

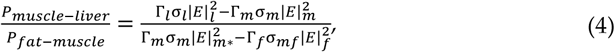

where subscript f denotes subcutaneous fat, and electric field strengths |E|_m_ and |E|_m_*are muscles adjacent to the liver and to the subcutaneous fat, respectively. The left side of the equation is from the measurements, and the right side is from the theory and the simulation. The left side of the equation is obtained by measuring the signal strengths of both boundaries. Acquired acoustic signal measurements are beamformed, and then signals corresponding to each boundary are calculated. Acoustic attenuation during the signal propagation is also compensated. In this equation, the unknowns are the subject’s liver tissue properties and the electric field strengths. The electric field distribution, even at other tissues, is affected by the electrical properties of the liver. Equation (4) can be rewritten as an explicit function of the liver fat content, ζ, by grouping all terms independent of ζ on the left-hand side:

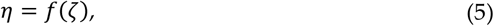

where η comprises the ratio of the measured signals and all other terms independent of ζ. The liver fat content can then be estimated by solving the following optimization problem:

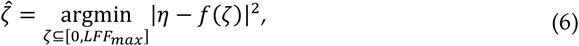

where LFF_max_ is the maximum expected liver fat fraction, which was assumed to be 40%.

## Appendix B

### Raw data

Table 5 contains the raw data set that was obtained during the feasibility study and used in this paper.

**Table 5.**
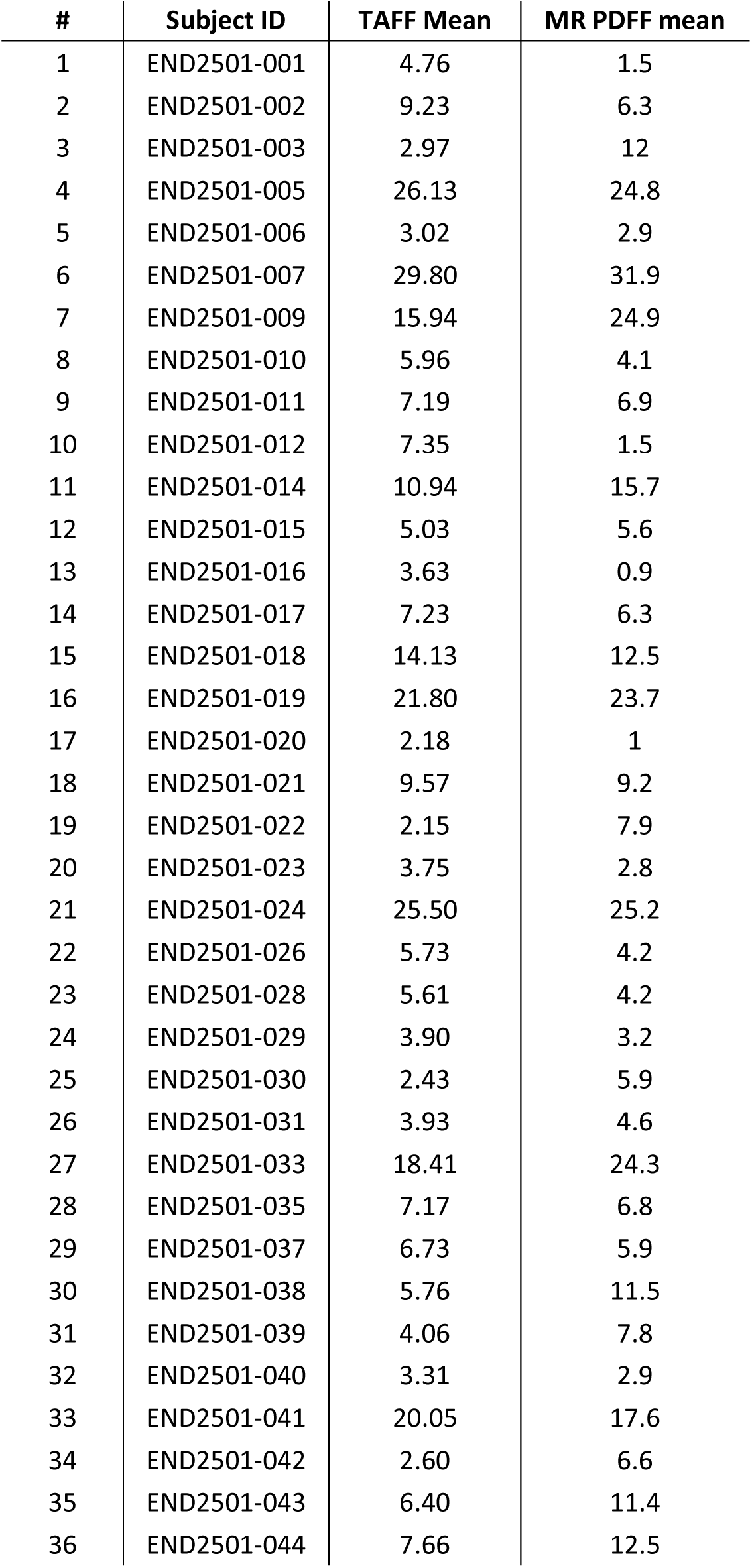
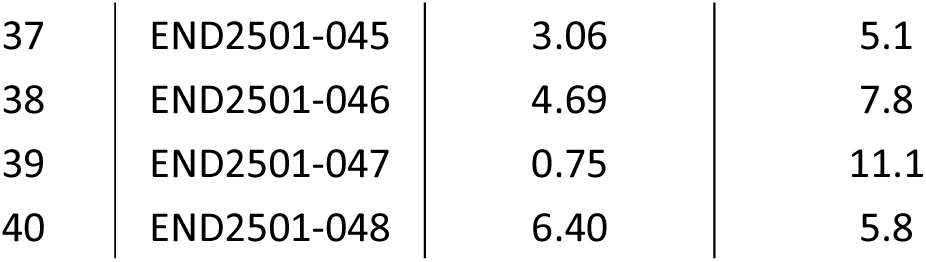
Raw data set from feasibility study:

| # | Subject ID | TAFF Mean | MR PDFF mean |
| --- | --- | --- | --- |
| 1 | END2501-001 | 4.76 | 1.5 |
| 2 | END2501-002 | 9.23 | 6.3 |
| 3 | END2501-003 | 2.97 | 12 |
| 4 | END2501-005 | 26.13 | 24.8 |
| 5 | END2501-006 | 3.02 | 2.9 |
| 6 | END2501-007 | 29.80 | 31.9 |
| 7 | END2501-009 | 15.94 | 24.9 |
| 8 | END2501-010 | 5.96 | 4.1 |
| 9 | END2501-011 | 7.19 | 6.9 |
| 10 | END2501-012 | 7.35 | 1.5 |
| 11 | END2501-014 | 10.94 | 15.7 |
| 12 | END2501-015 | 5.03 | 5.6 |
| 13 | END2501-016 | 3.63 | 0.9 |
| 14 | END2501-017 | 7.23 | 6.3 |
| 15 | END2501-018 | 14.13 | 12.5 |
| 16 | END2501-019 | 21.80 | 23.7 |
| 17 | END2501-020 | 2.18 | 1 |
| 18 | END2501-021 | 9.57 | 9.2 |
| 19 | END2501-022 | 2.15 | 7.9 |
| 20 | END2501-023 | 3.75 | 2.8 |
| 21 | END2501-024 | 25.50 | 25.2 |
| 22 | END2501-026 | 5.73 | 4.2 |
| 23 | END2501-028 | 5.61 | 4.2 |
| 24 | END2501-029 | 3.90 | 3.2 |
| 25 | END2501-030 | 2.43 | 5.9 |
| 26 | END2501-031 | 3.93 | 4.6 |
| 27 | END2501-033 | 18.41 | 24.3 |
| 28 | END2501-035 | 7.17 | 6.8 |
| 29 | END2501-037 | 6.73 | 5.9 |
| 30 | END2501-038 | 5.76 | 11.5 |
| 31 | END2501-039 | 4.06 | 7.8 |
| 32 | END2501-040 | 3.31 | 2.9 |
| 33 | END2501-041 | 20.05 | 17.6 |
| 34 | END2501-042 | 2.60 | 6.6 |
| 35 | END2501-043 | 6.40 | 11.4 |
| 36 | END2501-044 | 7.66 | 12.5 |
| 37 | END2501-045 | 3.06 | 5.1 |
| 38 | END2501-046 | 4.69 | 7.8 |
| 39 | END2501-047 | 0.75 | 11.1 |
| 40 | END2501-048 | 6.40 | 5.8 |

Table 6 contains the raw data set that was obtained during the R&R study and used in this paper.

**Table 6.** Raw data set from R&R study:

| Subject ID | User ID | Trial ID | TAFF |
| --- | --- | --- | --- |
| MCOM-021 | U1 | 1 | 3.76 |
| MCOM-021 | U2 | 1 | 3.84 |
| MCOM-021 | U1 | 2 | 5.1 |
| MCOM-021 | U2 | 2 | 7.6 |
| MCOM-022 | U1 | 1 | 12.28 |
| MCOM-022 | U2 | 1 | 11.86 |
| MCOM-022 | U1 | 2 | 11.18 |
| MCOM-022 | U2 | 2 | 7.92 |
| MCOM-023 | U1 | 1 | 8.1 |
| MCOM-023 | U2 | 1 | 12.48 |
| MCOM-023 | U1 | 2 | 9.58 |
| MCOM-023 | U2 | 2 | 10.22 |
| MCOM-024 | U1 | 1 | 3.58 |
| MCOM-024 | U2 | 1 | 5.64 |
| MCOM-024 | U1 | 2 | 3.5 |
| MCOM-024 | U2 | 2 | 7.02 |
| MCOM-025 | U1 | 1 | 15.36 |
| MCOM-025 | U2 | 1 | 8.16 |
| MCOM-025 | U1 | 2 | 15.34 |
| MCOM-025 | U2 | 2 | 11 |
| MCOM-026 | U1 | 1 | 7.16 |
| MCOM-026 | U2 | 1 | 7.16 |
| MCOM-026 | U1 | 2 | 6.68 |
| MCOM-026 | U2 | 2 | 7.36 |
| MCOM-027 | U1 | 1 | 20.34 |
| MCOM-027 | U2 | 1 | 13.32 |
| MCOM-027 | U1 | 2 | 9.42 |
| MCOM-027 | U2 | 2 | 23.12 |
| MCOM-028 | U1 | 1 | 20.02 |
| MCOM-028 | U2 | 1 | 11.44 |
| MCOM-028 | U1 | 2 | 7.98 |
| MCOM-028 | U2 | 2 | 22.5 |
| MCOM-029 | U1 | 1 | 5.52 |
| MCOM-029 | U2 | 1 | 7.3 |
| MCOM-029 | U1 | 2 | 5.16 |
| MCOM-029 | U2 | 2 | 10.36 |
| MCOM-030 | U1 | 1 | 10.46 |
| MCOM-030 | U2 | 1 | 12.34 |
| MCOM-030 | U1 | 2 | 10.48 |
| MCOM-030 | U2 | 2 | 9.92 |
| MCOM-031 | U1 | 1 | 15.94 |
| MCOM-031 | U2 | 1 | 9.4 |
| MCOM-031 | U1 | 2 | 13.22 |
| MCOM-031 | U2 | 2 | 12.76 |
| MCOM-032 | U1 | 1 | 13.96 |
| MCOM-032 | U2 | 1 | 8.7 |
| MCOM-032 | U1 | 2 | 12 |
| MCOM-032 | U2 | 2 | 13.36 |
| MCOM-033 | U1 | 1 | 20.88 |
| MCOM-033 | U2 | 1 | 18.02 |
| MCOM-033 | U1 | 2 | 14.14 |
| MCOM-033 | U2 | 2 | 19.26 |
| MCOM-034 | U1 | 1 | 31.6 |
| MCOM-034 | U2 | 1 | 26.96 |
| MCOM-034 | U1 | 2 | 30.46 |
| MCOM-034 | U2 | 2 | 34.4 |

## Appendix C

### Example of a beamformed thermoacoustic image

Figure 8 shows an example of beamformed image for a TAEUS scan along with an MRI and an US image. Tissue signals that are relevant to calculating the scan quality and estimating TAFF are marked with green circles. Thermoacoustic signals appear strongly at tissue boundaries. The presented ultrasound image was taken before the start of the thermoacoustic session, but the overlaid images show that there is a good agreement in tissue boundaries and thermoacoustic signals.

**Figure 8.**
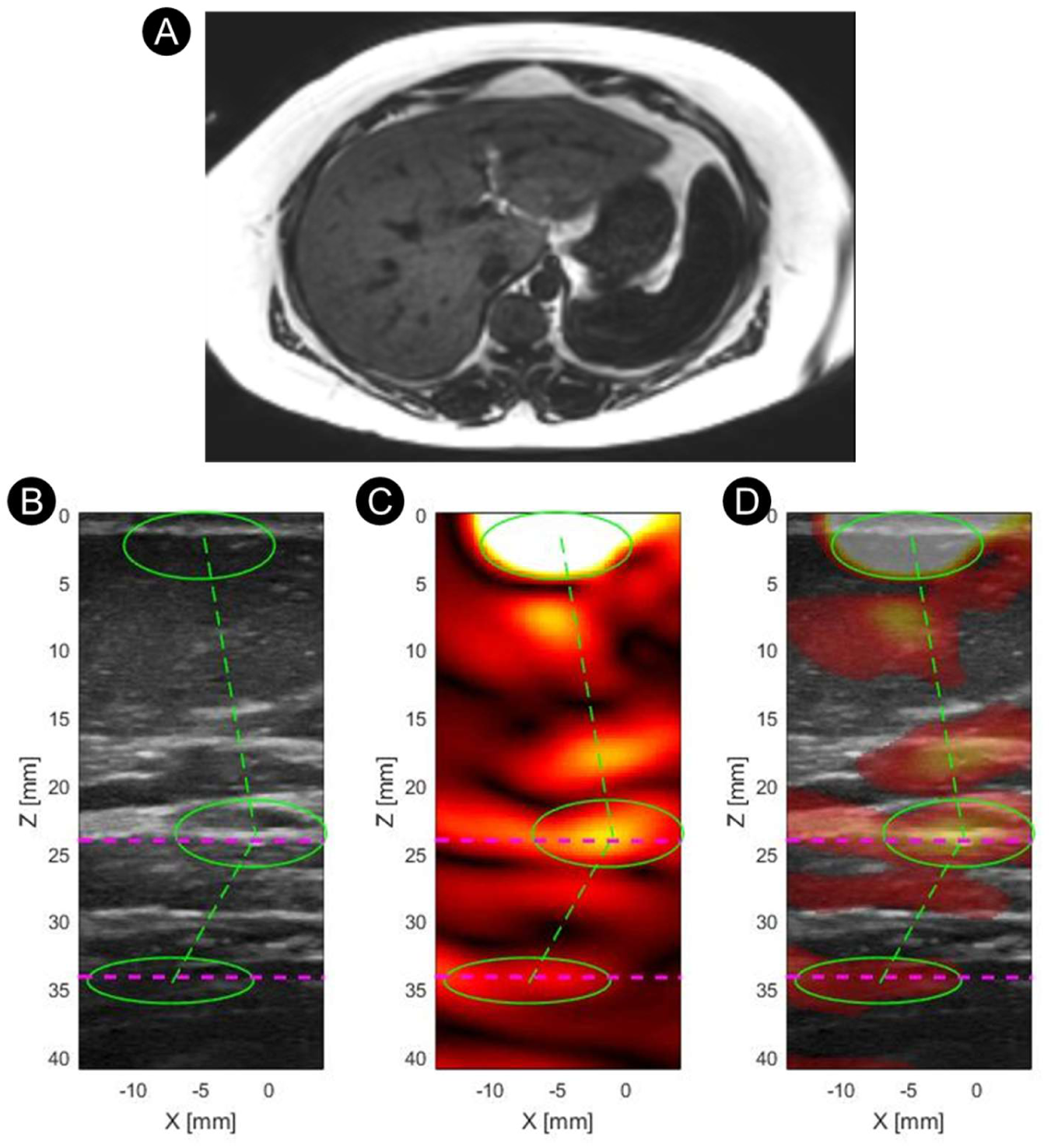
An example of a TAEUS beamformed image. (A) A MR image (mDIXON-Quant breath-hold (BH)) of the subject. (B) A linear US image at the scan location. (C) Beamformed TAEUS image (D) Beamformed TAEUS image overlaid on the US image. Green circles denote TAEUS signals from tissue boundaries relevant to the TAFF estimation: From top to bottom, probe-to-skin boundary, subcutaneous fat-to-muscle boundary, and muscle-to-liver boundary. Green dashed lines illustrate the spatial relationships among these boundary signals. Magenta dashed lines denote the tissue boundary locations identified during the ultrasound imaging.

**Disclaimer/Publisher’s Note:** The statements, opinions and data contained in all publications are solely those of the individual author(s) and contributor(s) and not of MDPI and/or the editor(s). MDPI and/or the editor(s) disclaim responsibility for any injury to people or property resulting from any ideas, methods, instructions or products referred to in the content.

